# Social participation and life satisfaction of peoples during the COVID-19 home confinement: the ECLB-COVID19 multicenter study

**DOI:** 10.1101/2020.05.05.20091066

**Authors:** Achraf Ammar, Hamdi Chtourou, Omar Boukhris, Khaled Trabelsi, Liwa Masmoudi, Michael Brach, Bassem Bouaziz, Ellen Bentlage, Daniella How, Mona Ahmed, Patrick Mueller, Notger Mueller, Asma Aloui, Omar Hammouda, Laisa Liane Paineiras-Domingos, Annemarie Braakman-jansen, Christian Wrede, Sophia Bastoni, Carlos Soares Pernambuco, Leonardo Mataruna, Morteza Taheri, Khadijeh Irandoust, Aïmen Khacharem, Nicola L Bragazzi, Karim Chamari, Stephen J Bailey, Nicholas T Bott, Faiez Gargouri, Lotfi Chaari, Hadj Batatia, Gamal Mohamed Ali, Osama Abdelkarim, Mohamed Jarraya, Kais El Abed, Nizar Souissi, Lisette Van Gemert-Pijnen, Bryan L Riemann, Laurel Riemann, Wassim Moalla, Jonathan Gómez-Raja, Monique Epstein, Robbert Sanderman, Sebastian Schulz, Achim Jerg, Ramzi Al-Horani, Taysir Mansi, Mohamed Jmail, Fernando Barbosa, Fernando Santos, Boštjan Šimunič, Rado Pišot, Donald Cowan, Andrea Gaggioli, Jordan M Glenn, Jürgen Steinacker, Tarak Driss, Anita Hoekelmann

## Abstract

**Background:** Public health recommendations and governmental measures during the COVID-19 pandemic have enforced numerous restrictions on daily living including social distancing, isolation and home confinement. While these measures are imperative to mitigate spreading of COVID-19, the impact of these restrictions on psychosocial health is undefined. Therefore, an international online survey was launched in April 2020 in seven languages to elucidate the behavioral and lifestyle consequences of COVID-19 restrictions. This report presents the preliminary results from the first thousand responders on social participation and life satisfaction.

**Methods:** Thirty-five research organisations from Europe, North-Africa, Western Asia and the Americas promoted the survey through their networks to the general society, in English, German, French, Arabic, Spanish, Portuguese, and Slovenian languages. Questions were presented in a differential format with questions related to responses “before” and “during” confinement conditions.

**Results:** 1047 replies (54% women) from Asia (36%), Africa (40%), Europe (21%) and other (3%) were included in the analysis. Preliminary findings revealed psychosocial strain during the enforced COVID-19 home confinement. In particular, large decreases in the amount of social activity through family (58%), friends/neighbors (44.9%) or entertainment (46.7%) were triggered by the enforced confinement. These negative effects on social participation were also associated with lower satisfaction (−30.5%) during the confinement period. Conversely, social contact score through digital technologies has significantly increased (p<0.001) during the confinement period with more individuals (24.8%) being socially connected through digital technology.

**Conclusion:** These preliminary findings elucidate the risk of psychosocial strain during the current home confinement period. Therefore, in order to mitigate the negative psychosocial effects of home confinement, implementation of national strategies focused on promoting social inclusion through technology-based solution is urgently needed.

## Introduction

Social participation and engagement requires the maintenance of a variety of social connections and relationships, as well as involvement in social and community activities.^1,2^ Examples of these activities include visiting and having contact with family and friends,^3^ belonging to religious groups,^4^ participating in occupational or social roles (e.g., volunteering for association or nonprofit organization;^5^), voting^6^ engagement in cultural and sports activities,^7^ and attending meetings.^8^ A good social participation boosts feeling of attachment, provides a consistent and coherent sense of identity, and enhances the sense of value, belonging and attachment to the individual’s community.^5^ In this context, Prilleltensky et al.^9^ reported that integration into community life and participation in social activities actively increases psychological and social wellbeing as well as an individual’s sense of belonging. In the same way, Smetana et al.^10^ showed social participation enhances self-efficacy and personal self-control in adolescents.

Termed “social health,” the enhancement of social participation is one of the important targets for health professionals.^11^ As indicated by Levasseur et al.^12^, social participation is related to mortality, morbidity, and life quality. The World Health Organization’ (WHO) recommends that particular attention should be given to social participation, especially for elderly as they spend less time in structured employment.^13^ Also, social participation plays an important positive role in personal wellbeing (e.g., life satisfaction)^14^ and social wellbeing^5^ for adolescents and adults. On the other hand, participating in personal leisure activities (a form of social participation) is of high importance for physical health, mental health, and improved quality of life.^15^

Social participation and life satisfaction are strongly related.^5^ Life satisfaction is defined as the estimation of life quality based on an individual’s preferences and satisfaction in these domains.^16^ For social wellbeing, life satisfaction is of crucial importance. Indeed, it has been reported that life satisfaction is associated with psychiatric disorders (e.g., depressive disorders) and suicidal ideation.^16^

A novel coronavirus, named SARS-CoV-2 or COVID-19, was detected in Hubei, China in December 2019. As of this publication (April 11,2020), COVID-19 was reported in more than 200 countries, affecting more than 2,350,000 peoples and resulting in more than 160,000 deaths. The COVID-19, which was declared a global pandemic on March 11^th^, 2020, is a serious challenge, facing all societies. To decelerate its rapid transmission, social confinement remains the best non-pharmacological solution and as a result, many countries have imposed stringent social distancing measures. While quarantine has been utilised previously to combat infectious diseases (e.g., cholera, SARS, Ebola), the level of confinement applied to the global population is the most severe in history.

Although it is the most effective solution to slow the spread of infectious disease, home confinement can also have negative effects on social participation and life satisfaction. This crisis (i.e., COVID-19 spread and the associated confinement) may also be associated with sensations of loneliness, grief, and loss of life satisfaction. Possible relationship changes with family and friends, as well as mitigated participation in community life are expected. The purpose of this investigation is to examine the effects of home confinement on social participation and life satisfaction. Results of this study could provide conclusions about confinement related social participation and life satisfaction changes; the ultimate goal being to highlight the importance of setting up programs to support individuals as they go through this crisis.

## Methods

We report findings on the first 1047 replies to an international online-survey on mental health and multi-dimension lifestyle behaviors during home confinement (ECLB-COVID19). ECLB-COVID19 was opened on April 1, 2020, tested by the project’s steering group for a period of 1 week, before starting to spread it worldwide on April 6, 2020. Forty-one research organizations from Europe, North-Africa, Western Asia and the Americas promoted dissemination and administration of the survey. ECLB-COVID19 was administered in English, German, French, Arabic, Spanish, Portuguese, and Slovenian languages. The survey included sixty-four questions on health, mental wellbeing, mood, life satisfaction and multidimension lifestyle behaviors (physical activity, diet, social participation, sleep, technology-use, need of psychosocial support). All questions were presented in a differential format, to be answered directly in sequence regarding “before” and “during” confinement conditions. The study was conducted according to the Declaration of Helsinki. The protocol and the consent form were fully approved (identification code: 62/20) by the Otto von Guericke University Ethics Committee.

## Study design and study sample

The ECLB-COVID19 electronic survey was designed by a steering group of multidisciplinary scientists and academics (i.e., human science, sport science, neuropsychology, computer science) at the University of Magdeburg (principal investigator), the University of Sfax, the University of Münster, and the University of Paris-Nanterre; development followed an initial structured review of the literature. The survey was then reviewed and edited by Over 50 colleagues and experts worldwide. The survey was uploaded and shared online via the Google platform. A link to the electronic survey was distributed worldwide by consortium colleagues via a range of methods: invitation via e-mails, shared in consortium’s faculties official pages, ResearchGate™, LinkedIn™ and other social media platforms such as Facebook™, WhatsApp™ and Twitter™. Public were also involved in the dissemination plans of our research through the promotion of the ECLB-COVID19 survey in their networks. The survey included an introductory page describing the background and the aims of the survey, the consortium, ethics information for participants and the option to choose one of seven available languages (English, German, French, Arabic, Spanish, Portuguese, and Slovenian). The present study focusses on the first thousand responses (i.e., 1047 participants), which were reached on April 11, 2020, approximately one-week after the survey began. This survey was open for all people worldwide aged 18 years or older. People with cognitive decline were excluded.

## Data privacy and consent of participation

During the informed consent process, survey participants were assured all data would be used only for research purposes. Participants’ answers are anonymous and confidential according to Google’s privacy policy (https://policies.google.com/privacy?hl=en). Participants don’t have to mention their names or contact information. In addition, participant can stop participating in the study and can leave the questionnaire at any stage before the submission process and their responses will not be saved. Response will be saved only by clicking on “submit” button. By completing the survey, participants are acknowledging the above approval form and are consenting to voluntarily participate in this anonymous study. Participants have been requested to be honest in their responses.

## Survey questionnaires

The ECLB-COVID19 is a multi-country electronic survey designed to assess change in multiple lifestyle behaviors during the COVID-19 outbreak. Therefore, a collection of validated and/or crisis-oriented briefs questionnaires were included. These questionnaires assess mental wellbeing (Short Warwick-Edinburgh Mental Well-being Scale (SWEMWBS)^17^), mood and feeling (Short Mood and Feelings Questionnaire (SMFQ)^18^), life satisfaction (Short Life Satisfaction Questionnaire for Lockdowns (SLSQL), social participation (Short Social Participation Questionnaire for Lockdowns (SSPQL)), physical activity (International Physical Activity Questionnaire Short Form (IPAQ-SF)^19,20^), diet behaviors (Short Diet Behaviors Questionnaire for Lockdowns (SDBQL)), sleep quality (Pittsburgh Sleep Quality Index (PSQI)^21^) and some key questions assessing the technology-use behaviors (Short Technology-use Behaviors Questionnaire for Lockdowns (STBQL)), demographic information and the need of psychosocial support. Reliability of the shortened and/or newly adopted questionnaires was tested by the project steering group through piloting, prior to survey administration. These brief crisis-oriented questionnaires showed good to excellent test-retest reliability coefficients (r = 0.84-0.96). A multi-language validated version already existed for the majority of these questionnaires and/or questions. However, for questionnaires that did not already exist in multi-language versions, we followed the procedure of translation and backtranslation, with an additional review for all language versions from the international scientists of our consortium. As a result, a total number of sixty-for items were included in the ECLB-COVID19 online survey in a differential format; that is, each item or question requested two answers, one regarding the period before and the other regarding the period during confinement. Thus, the participants were guided to compare the situations. Given the large number of questions included, the present paper focuses on newly developed SLSQL and SSPQL as brief crisis-oriented tools.

### Short Social Participation Questionnaire-Lockdowns (SSPQL)

The present Short Social Participation Questionnaire-Lockdowns (SSPQ-L) is a crisis-oriented short modified questionnaire to assess diet behavior before and during a lockdown period. The SDBQ-L is based on the eighteen items of the SPQ. The original SPQ items aim to ask respondents to indicate how regularly they had undertaken each activity in the last 12 months. From questions 1 to 12, participant could choose one of the six response categories: “Never”, “Rarely”, “A few times a year”, “Monthly”, “A few times a month”, and “Once a week or more”. The remaining four items requested a binary “Yes” or “No” response regarding participation in community groups in the last 12 months.^22^ Given that we are assessing social participation before and during the home confinement, which is a short period (days to months), we adapted the response categories and shortened the number of questionnaires by combining similar questions (e.g., Q1 and 2; Q2 and Q3; Q12 and Q14), while adding one more question about the use of phone calls for communication. Accordingly, the final SSPQ-L includes 14 items with five response categories (i.e., “Never”=1 point; “Rarely”=2 points; “Sometimes”=3 points; “Often”=4 points and “All times”=5 points) for the 10 first items and “Yes”=5 points / “No”=1 point response categories for the four remaining items. Total scores of this questionnaire correspond to the sum of the scored points in the 14 questions. The total score for the SSPQ-L is from “14” to “70”, where “14” indicate that participant has “never” being socially active; a score between “15” and “28” indicate that participant has “rarely” being socially active, a score between “29” and “42” indicate that participant is “sometimes” socially active, a score between “43” and “56” indicate that participant is “often” socially active, and a score between “57” and “70” indicate that participant is at “all times” socially active.

### Short Life Satisfaction Questionnaire for Lockdowns (SLSQL)

The present Short Life Satisfaction Questionnaire-Lockdowns (SLSQL) is a crisis-oriented short modified questionnaire to assess satisfaction with the respondent’s life as a whole before and during the confinement period. The SLSQL is the short version of the Satisfaction With Life Scale (SWLS)’s five items.^23^ Three questions from the SWLS questionnaire, that showed to be related to emotional well-being, were included to allow an individuals’ conscious evaluative judgment of participant life by using the person’s own criteria. Using the 1 -7 scale below, Participants indicate their agreement with each of the three included items (Strongly agree=7; Agree=6; Slightly agree=5; Neither agree nor disagree=4; Slightly disagree=3; Disagree=2; Strongly disagree=1). Total score of this questionnaire, correspond to the sum of the scored points in the 3 questions. The total score for the SLSQ-L is from “3” to “21”, where “3” indicate that participant is “Extremely dissatisfied”, “4-6” indicate that participant is “dissatisfied”, “7-9” indicate that participant is “Slightly dissatisfied”, “10-12” indicate that participant is “Neutral”, “13-15” indicate that participant is “Slightly satisfied”, “16-18” indicate that participant is “satisfied”, and “19-21” indicate that participant is “Extremely satisfied”.

## Data Analysis

Descriptive statistics were used to define the proportion of responses for each question and the total distribution of the total score of each questionnaire. All statistical analyses were performed using the commercial statistical software STATISTICA (StatSoft, Paris, France, version 10.0). Normality of the data distribution was confirmed using the Shapiro-Wilks-W-test. Values were computed and reported as mean ± SD (standard deviation). Paired samples t-tests were used to assess significant difference in total scored responses between “before” and “during” confinement period. Effect size (Cohen’s d) was calculated to determine the magnitude of the change score and interpreted using the following criteria: 0.2 (small), 0.5 (moderate), and 0.8 (large).^24^ Pearson product-moment correlation tests were used to assess possible relationships between the “before-after” Δ of the assessed multidimension total scores. Statistical significance was identified at p<0.05.

## Results

### Sample description

1047 participants were included in the preliminary sample. Overall, 54% of the sample were women and 46% were men. Geographical breakdowns were from Asian (36%, mostly from Western Asia), African (40%, mostly from North Africa), European (21%) and other (3%) countries. Age, health status, employment status, level of education, and marital status are presented in Table 1.

**Table 1:**
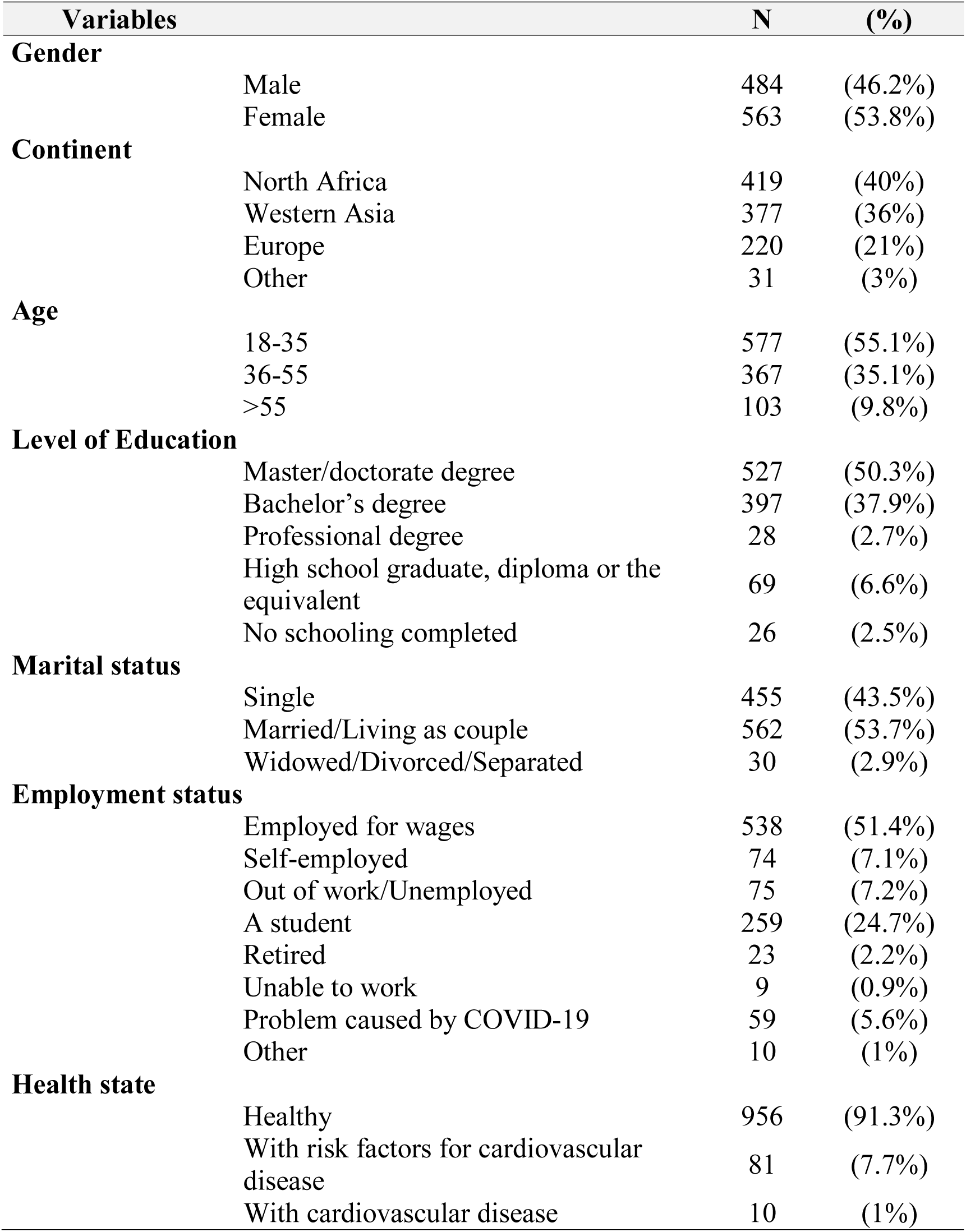
Demographic characteristics of the participants (N = 1047)

### Short Social Participation Questionnaire for Lockdowns (SSPQL)

Change in social participation score from “before” to “during” confinement period in response to SSPQL assessment tool are presented in table 2. Statistical analysis showed the total score of SSPQL decreased significantly by 42% “during” compared to “before” home confinement (t=69.19 p<0.001, d=2.14). This significant decrease was observed in the score recorded by each question (1 to 14). Particularly, the recorded score in social participation through family, neighbors, friend or church or religious activities (Q1-Q3) were lower at “during” compared to “before” confinement with |Δ%| ranged from 56% to 59% (34.9 ≤ t ≤ 54.9; P < 0.001, 1.07 ≤ d ≤ 1.7). Similarly, questions related to going to entertainment places (Q6-Q9), participating in community group activities (Q11-Q14) or doing other form of activities (Q10) that provide social contact showed lower scores at “during” compared to “before” confinement with |Δ%| ranged from 12.1% to 68.4% (5.95 ≤ t ≤ 65.77; P< 0.001, 0.18 ≤ d ≤ 2.03). However, scores related to social contact through technology-use behaviors (Q4 and Q5) increased at “during” compared to “before” the confinement period with |Δ%| ranged between 5.7% and 10.2% (7.20 ≤ t ≤ 65.77; P < 0.001, 0.22 ≤ d ≤ 2.03.

**Table 2.**
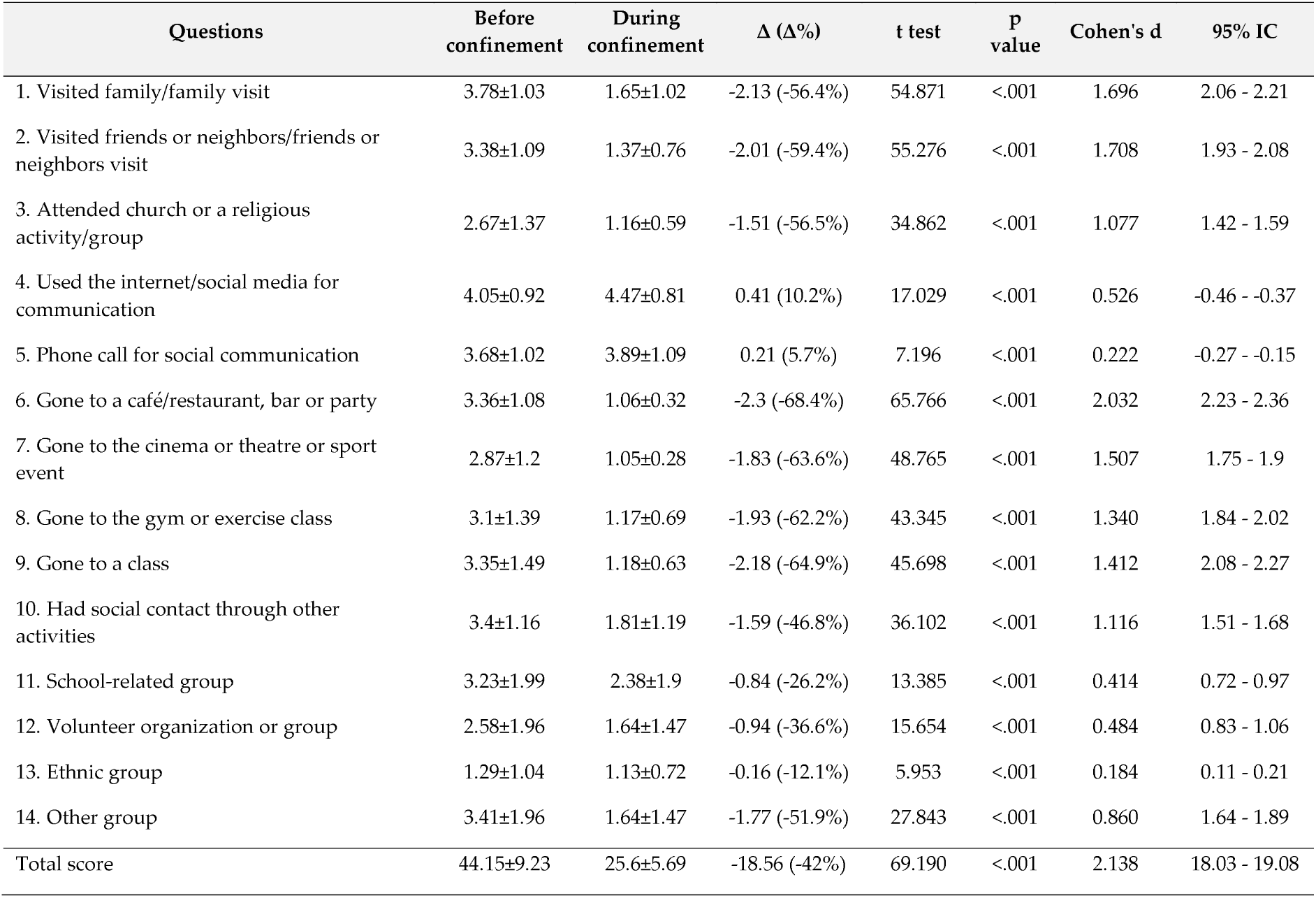
Responses to Short Social Participation Questionnaire-Lockdowns before and during home.

| Questions | Before<br>confinement | During<br>confinement | $\Delta$ ($\Delta\%$) | t test | P<br>value | Cohen's d | 95% IC |
| --- | --- | --- | --- | --- | --- | --- | --- |
| 1. Visited family/family visit | 3.78±1.03 | 1.65±1.02 | -2.13 (-56.4%) | 54.871 | <.001 | 1.696 | 2.06 - 2.21 |
| 2. Visited friends or neighbors/friends or neighbors visit | 3.38±1.09 | 1.37±0.76 | -2.01 (-59.4%) | 55.276 | <.001 | 1.708 | 1.93 - 2.08 |
| 3. Attended church or a religious activity/group | 2.67±1.37 | 1.16±0.59 | -1.51 (-56.5%) | 34.862 | <.001 | 1.077 | 1.42 - 1.59 |
| 4. Used the internet/social media for communication | 4.05±0.92 | 4.47±0.81 | 0.41 (10.2%) | 17.029 | <.001 | 0.526 | -0.46 - -0.37 |
| 5. Phone call for social communication | 3.68±1.02 | 3.89±1.09 | 0.21 (5.7%) | 7.196 | <.001 | 0.222 | -0.27 - -0.15 |
| 6. Gone to a café/restaurant, bar or party | 3.36±1.08 | 1.06±0.32 | -2.3 (-68.4%) | 65.766 | <.001 | 2.032 | 2.23 - 2.36 |
| 7. Gone to the cinema or theatre or sport event | 2.87±1.2 | 1.05±0.28 | -1.83 (-63.6%) | 48.765 | <.001 | 1.507 | 1.75 - 1.9 |
| 8. Gone to the gym or exercise class | 3.1±1.39 | 1.17±0.69 | -1.93 (-62.2%) | 43.345 | <.001 | 1.340 | 1.84 - 2.02 |
| 9. Gone to a class | 3.35±1.49 | 1.18±0.63 | -2.18 (-64.9%) | 45.698 | <.001 | 1.412 | 2.08 - 2.27 |
| 10. Had social contact through other activities | 3.4±1.16 | 1.81±1.19 | -1.59 (-46.8%) | 36.102 | <.001 | 1.116 | 1.51 - 1.68 |
| 11. School-related group | 3.23±1.99 | 2.38±1.9 | -0.84 (-26.2%) | 13.385 | <.001 | 0.414 | 0.72 - 0.97 |
| 12. Volunteer organization or group | 2.58±1.96 | 1.64±1.47 | -0.94 (-36.6%) | 15.654 | <.001 | 0.484 | 0.83 - 1.06 |
| 13. Ethnic group | 1.29±1.04 | 1.13±0.72 | -0.16 (-12.1%) | 5.953 | <.001 | 0.184 | 0.11 - 0.21 |
| 14. Other group | 3.41±1.96 | 1.64±1.47 | -1.77 (-51.9%) | 27.843 | <.001 | 0.860 | 1.64 - 1.89 |
| Total score | 44.15±9.23 | 25.6±5.69 | -18.56 (-42%) | 69.190 | <.001 | 2.138 | 18.03 - 19.08 |

### Short Life Satisfaction Questionnaire-Lockdowns

Change in life satisfaction score from “before” to “during” confinement period in response to SLSQL are presented in table 3. Statistical analysis showed that the total score of SLSQL decreased significantly by 16% “during” compared to “before” home confinement (t=21.05, p<0.001, d=0.65). This significant decrease was observed in the score recorded by the three included questions (Q1-Q3). Particularly, in response to the direct (Q3) and indirect (Q1-Q2) questions related to life satisfaction lower scores were recorded at “during” compared to “before” confinement with |Δ%| ranged from 14% to 18% (17.6 ≤ t ≤ 19.11; P < 0.001, 0.59 ≤ d ≤ 0.79).

**Table 3.** Responses related to Short Life Satisfaction Questionnaire-Lockdowns before and during home confinement.

| | Before<br>confinement | During<br>confinement | $\Delta$ ($\Delta\%$) | t test | p<br>value | Cohen's<br>d | 95% IC |
| --- | --- | --- | --- | --- | --- | --- | --- |
| 1. In most ways my life is close to my ideal | 4.81±1.62 | 3.93±1.71 | -0.88 (-18.2%) | 19.119 | <.001 | 0.591 | 0.79 - 0.97 |
| 2. So far I have gotten the important things I want in life | 4.67±1.7 | 4.0±1.81 | -0.67 (-14.4%) | 16.212 | <.001 | 0.501 | 0.59 - 0.75 |
| 3. I am satisfied with my life | 5.29±1.56 | 4.49±1.82 | -0.79 (-15%) | 17.560 | <.001 | 0.543 | 0.71 - 0.88 |
| Tot score | 14.77±4.32 | 12.42±4.67 | -2.34 (-15.9%) | 21.050 | <.001 | 0.651 | 2.12 - 2.56 |

## Discussion

To contain COVID-19 transmission, policymakers in many countries have considered implementing restrictive measures. Understanding the psychosocial implications of these measures will allow for better-informed decisions. The present study aims to provide insight into the effect of home-confinement on life satisfaction and social participation, based on data extracted from the first thousand multi-country responses. Preliminary results from 1047 participants (54% female; 36%, from Western Asia, 40% from North Africa, 21% from European and 3% from other countries) showed COVID-19 home confinement has a negative effect on social participation and life satisfaction. Total score in the social participation questionnaire decreased by 42% with more socially (+71.15%, Never-Rarely socially active) inactive individuals “during” compared to “before” the confinement period. Similarly, total score in life satisfaction questionnaires decreased by 16% with more people feeling dissatisfied (extremely-slightly) (+16.5%) “during” compared to “before” the confinement period. During similar pandemic crises (2002–2004 SARS outbreak), previous research revealed several negative effects of quarantine measures on social participation and were associated with reductions in individual wellbeing.^25,26^ These negative effects have also been reported in a recent COVI-19 series highlighting the fact that people in quarantine report a greater symptom of psychological distress; furthermore, some of these symptoms appear to stay long after the quarantine period ends.^27^ Similarly, results from Chinese studies indicate the COVID-19’s resultant social distancing measures engendered less life satisfaction and more distress. With significant negative effects of the current COVID-19 pandemic on social participation and life satisfaction scores, the present findings support these previous reports, elucidating the risk of psychosocial strain during the current home confinement period.

Particularly, the recorded total score during the home confinement was about 26pts (vs. 44pts before confinement), meaning that participants are rarely engaging in social activities, with a higher risk of social exclusion. This could be explained by social restrictions and reduced mobility imposed by governmental entities to contain the spread of the virus.^29^ Present findings indicate the 71.15% reduction in total social participation score was largely due to the decrease in social participation through family-visit activity (58%), with less individuals reporting regular (often/all times) visits to their family during compared to before the confinement period (7.2% vs. 65.2%). Social participation through entertainment activities or neighbors/friend visits recorded the second large decrease (44.9% to 46.7%), with the proportion of people declaring to regularly visit their neighbors/friends or regularly go to coffee shops/restaurants/parties decreased from more than 47% at before confinement to between 1% and 3% during confinement period. Of note is that younger populations showed a very large decrease in social participation through class participation, gym, or exercise activities (40% to 53%). The present widespread social isolation imposed by COVID-19 could induce a detrimental effect on mental health. Indeed, according to one study evaluating 1006 Italians under COVID-19 quarantine, longitudinal forced isolation increased depression, unworthiness, alienation, and helplessness.^29^ In addition, worse health conditions, as well as distress, were reported by adults who were not working in China.^28^

The present findings confirm the causal relationship between social participation and psychological health, showing a significant positive correlation between the total score recorded for social participation and life satisfaction (p<0.001 and r=0.23). Additionally, it was revealed that social distancing during home confinement was associated with less satisfied persons (−30.5%). Indeed, before the confinement more than 60 % agreed to being satisfied with their life, while during the confinement only 30% reported agreeing to this statement. Similarly, total scores of life satisfaction moved from “Slightly satisfied” (i.e., before home confinement) to “Neutral” (i.e., during home confinement). This close relationship between social distancing and life dissatisfaction may be due to the fact that socially distancing yourself from someone to which you are emotionally attached is difficult and can result in life dissatisfaction. Therefore, to keep an acceptable level of life satisfaction, it is important to stay connected while staying away. As our study indicates, social participation through family, friends and neighbors was most negatively affected by confinement, revealing the importance of staying in touch with friends and family to keep an acceptable level of life satisfaction.

The present findings also showed that social participation through internet, social media, phone calls etc. has increased from before to during home confinement with more individuals (24.8%) declaring the use of digital-facilities to stay socially connected during the home confinement period (61.5% vs. 36.7%). As participants demonstrated a greater use of technology during the confinement period, this medium may provide an avenue to foster social communication, thereby mitigating life dissatisfaction. Information and communications technology (ICT) such as video chat, social media, social platform, gamification, mhealth, interactive coach etc. can be therefore suggested to stay connected while staying away.

## Conclusions

The results from the first 1047 replies to the ECLB-COVID19 survey reveal a psychosocial strain during home confinement. In particular, a large decrease in social participation, triggered by enforced home confinement, was associated with lower satisfaction levels. Conversely, social contact through digital technology has increased during the confinement period. Therefore, in order to mitigate the negative psychosocial effects of home confinement, implementation of national strategies to promote social inclusion through ICT-based solutions are urgently needed. Additionally, given that present findings are based on data from heterogenic populations with no criteria-based subsamples analysis, further research is warranted to identify subpopulations that might be more affected by COVID-19 confinement measures. Identification of such populations would allow for better informed and more targeted mitigation strategies.

## Data Availability

Data are available by the correspondnat author upon raisonable request

## Acknowledgements

We thank our consortium’s colleagues who provided insight and expertise that greatly assisted the research. We thank all colleagues and peoples who believed on this initiative and helped to distribute the anonymous survey worldwide. We are also immensely grateful to all participants who #StayHome & #BoostResearch by voluntarily taken the #ECLB-COVID19 survey.

